# Strong off-target antibody reactivity to malarial antigens induced by RTS,S/AS01E vaccination is associated with increased protection

**DOI:** 10.1101/2021.12.23.21268281

**Authors:** Dídac Macià, Joseph J. Campo, Gemma Moncunill, Chenjerai Jairoce, Augusto J. Nhabomba, Maximilian Mpina, Hermann Sorgho, David Dosoo, Ousmane Traore, Kwadwo Asamoah Kusi, Nana Aba Williams, Arlo Randall, Hèctor Sanz, Clarissa Valim, Kwaku Poku Asante, Seth Owusu-Agyei, Halidou Tinto, Selidji Todagbe Agnandji, Simon Kariuki, Ben Gyan, Claudia Daubenberger, Benjamin Mordmüller, Paula Petrone, Carlota Dobaño

**Affiliations:** ISGlobal, Hospital Clínic - Universitat de Barcelona, Rosselló 153, 08036 Barcelona, Catalonia, Spain; Antigen Discovery, Inc (ADI), 1 Technology Dr., STE E309, Irvine, CA, USA; CIBER de Enfermedades Infecciosas, Spain; Centro de Investigação em Saúde de Manhiça (CISM), Rua 12, Cambeve, Vila de Manhiça, CP 1929, Maputo, Mozambique; Ifakara Health Institute. Bagamoyo Research and Training Centre. P.O. Box 74 Bagamoyo, Tanzania; Swiss Tropical and Public Health Institute, Socinstrasse 57, 4002 Basel, Switzerland; University of Basel, Petersplatz 1, 4001 Basel, Switzerland; Unité de Recherche Clinique de Nanoro, Institut de Recherche en Sciences de la Santé, Nanoro, BP 218, Burkina Faso; Department of Electron Microscopy & Histopathology, NMIMR, College of Health Sciences, University of Ghana, Legon; Kintampo Health Research Centre, P.O. Box 200, Kintampo, Brong-Ahafo, Ghana; Department of Immunology and Infectious Diseases, Harvard T.H. Chen School of Public Health. 675 Huntington Ave., Boston MA 02115, USA; Centre de Recherches Médicales de Lambaréné (CERMEL), BP 242, Lambaréné, Gabon; Kenya Medical Research Institute/Centre for Global Health, Kisumu, Siaya, P.O. Box 54840 00200 Nairobi, Kenya; Noguchi Memorial Institute for Medical Research, University of Ghana, P.O. Box LG 581 Legon Ghana; Institute of Tropical Medicine and German Center for Infection Research, University of Tübingen, Wilhelmstraße 27 D-72074 Tübingen, Germany

## Abstract

The RTS,S/AS01E vaccine targets the circumsporozoite protein (CSP) of the *Plasmodium falciparum* parasite. Using protein microarrays, levels of IgG to 1,000 *P. falciparum* antigens were measured in 2,138 infants (age 6-12 weeks) and children (age 5-17 months) from 6 African sites of the phase 3 trial, sampled before and at four longitudinal visits after vaccination. One month post-vaccination, IgG responses to 17% of all probed antigens showed differences between RTS,S/AS01E and comparator vaccination groups, whereas no pre-vaccination differences were found. A small subset of antigens presented IgG levels reaching 4- to 8-fold increases in the RTS,S/AS01E group, comparable in magnitude to anti-CSP IgG levels (∼11-fold increase). They were strongly cross-correlated and correlated with anti-CSP levels, waning similarly over time and re-increasing with the booster dose. Such an intriguing phenomenon may be due to cross-reactivity of anti-CSP antibodies with these antigens. RTS,S/AS01E vaccinees with strong off-target IgG responses had an estimated lower clinical malaria incidence after adjusting for age group, site and post-vaccination anti-CSP levels. RTS,S/AS01E-induced IgG may bind strongly not only to CSP, but to unrelated malaria antigens, and this seems to either confer, or at least be a marker of, increased protection from clinical malaria.

## Introduction

The recombinant protein in adjuvant subunit vaccine RTS,S/AS01E (Mosquirix) is the first to be recommended by the WHO for use in African children to prevent malaria, following the completion of a phase 3 trial for licensure and a pilot implementation study (WHO, 2021). This vaccine targets *Plasmodium falciparum*, the deadliest of malaria species in humans, a protozoan parasite expressing over 5,300 proteins, of which hundreds are located near the parasite surface at some stage of its lifecycle, thereby presenting numerous targets for human host antibodies (Abs). The vaccine contains a part of the circumsporozoite (CSP) sequence, a crucial protein on the sporozoite surface which targets *P. falciparum* parasites before they reach the liver, and it is co-expressed with hepatitis B surface antigen (HBsAg). Vaccine efficacy (VE) was assessed in the phase 3 trial between 2009 and 2014 in 11 sites in Africa, yielding estimates of 55.8% in children (The RTS,S Partnership, 2011) and 31.3% in infants over 1 year of follow-up (The RTS,S Partnership, 2012), yet rapidly decreasing in the following years (RTS,S and Partnership, 2015). Our present study is a nested immunology study within the phase 3 clinical trial.

Immunogenicity studies have established a dose–response relationship between post-vaccination anti-CSP Ab levels and VE across multiple trial sites (Dobaño et al., 2019, White et al., 2014), with Ab waning curves over succeeding months closely coinciding with VE decline. Likewise, in other subunit vaccines, circulating Ab levels to the target protein are frequently the best measurable correlate of VE. By contrast, monitoring Ab responses to proteins from the same pathogen that are not included in the vaccine (off-target) is less common and, when pursued, the objective usually resides in describing the complex interplay of vaccine-induced and naturally acquired immunity against the same pathogen (Campo et al., 2015, Dobaño et al., 2019a, Dobaño et al., 2019b). After RTS,S/AS01E vaccination in endemic settings, a number of off-target Ab levels, mainly markers of exposure, were lower in RTS,S/AS01E vaccinees in the months following vaccination (Bejon et al., 2011; Campo et al., 2011; Campo et al., 2015). These results indicate that acquisition of natural immunity is reduced, or more probably delayed (White et al., 2015), as a consequence of the partial protection conferred by the vaccine.

Unexpectedly, a small group of vaccine off-target Abs has also been found that appeared to be increased in vaccinated children. In a panel of more than 800 proteins tested in samples taken 6 months after primary vaccination from a Mozambican phase 2b trial, a large number of Abs showed expected decreases in RTS,S/AS01E vaccinees, but a much smaller number of Abs were increased, which became a majority when the comparison was restricted to vaccinees with no reported clinical malaria cases during follow-up (Campo et al., 2015). More recently, a panel of 40 proteins probed in plasmas sampled 1 month after vaccination from the phase 3 clinical trial showed a small group of antigens (Ags) with increased Ab levels in the RTS,S group (Dobaño et al., 2019a, Dobaño et al., 2019b). More surprisingly, some of these off-target Ab increases induced only one month following vaccination were associated with greater protection. In light of this, we then speculated that asymptomatic malaria infections could still take place in RTS,S vaccinees and that these low-level infections could account for an accelerated acquisition of natural immunity. We also considered the possibility of Ab cross-reactivity even when no clear sequence similarities were found.

In vaccinology, cross-reactivity has mainly been studied between key Ags from different pathogen strains or species (heterologous or between-pathogen cross-reactivity) often motivated by the cross-protection that may result thereof (Benn et al., 2013; Stojanovic et al., 2020; Vojtek et al., 2019). By contrast, little is known about cross-reactivity between Ags expressed in the same organism (within-pathogen cross-reactivity) and the potential reinforced protection that may result. However, evidence exists that a degree of cross-reactivity between epitopes of different malarial proteins expressed at different life-cycle stages is possible (Hope et al., 1984; Ardeshir et al., 1990) and seems to be mainly driven by immunodominant low-complexity repeat structures characteristic of the malaria parasite, such as those in CSP (Feng et al., 2006; Hou et al., 2020). The advent of high throughput immunology, capable of screening a specific Ab response against nearly all Ags expressed in the same pathogen, will likely challenge assumptions of within-pathogen cross-reactivity.

The aim of this study was to address the phenomenon of vaccine-induced off-target Ab reactivity occurring immediately after vaccination and through long-term follow-up with booster administration using an unbiased and comprehensive panel of Ags. We used this rich dataset to identify direct causal effects of RTS,S/AS01E vaccination on off-target Abs, which were expected to be highest after vaccination and wane over follow-up months similar to anti-CSP Ab levels, as opposed to indirect effects mediated by differential natural acquired immunity, which were expected to build up over time and be negatively associated with vaccination due to reduced parasite exposure.

Our analyses of vaccine off-target Ab reactivity covered a panel of 1,000 *P. falciparum* antigenic proteins or fragments representing 762 unique protein-coding genes (14% of the 3D7 strain proteome), and allowed us to assess at a large scale whether RTS,S/AS01E-induced off-target Ab reactivity to different Ags is independent of one another or is organised in signatures. We also investigated its association with age, exposure and other epidemiologic variables. Finally, we assessed whether the presence and intensity of off-target Ab responses immediately following vaccination is a correlate of VE even after adjusting for anti-CSP levels.

## Results

### IgG against many *P. falciparum* proteins are altered after primary vaccination with a small group of proteins showing high increases

Prior to vaccination (study month 0, “M0”), Ab levels exhibited no significant differences to any probed Ags in the microarray between the RTS,S and comparator groups (***Figure 1A***, left). This result indicated that the study subsample preserved the randomization of the clinical trial vaccination groups and there existed no differences in baseline immunological profiles prior to intervention. Only a month after the primary vaccination concluded (M3), there were significant differences between vaccination groups in 17% of all probed Ags (***Figure 1A***, right). Some of these Ags elicited surprisingly strong mean Ab level increases, approaching that of CSP (11.5-fold increase in the RTS,S over the comparator group, CI [11.0-12.3]). Ranking them in decreasing order and only including the largest increases (>2-fold), we encountered: a claudin-like apicomplexan microneme protein (“CLAMP”, 7.8, CI [7.2-8.5]), a glycogen synthase kinase (“GSK3”, 5.4, CI [4.9,5.9]), a RING zinc finger protein (“RZnFing”, 4.6, CI [4.2,5.0]), the Merozoite Surface Protein 5 (“MSP5”, 4.0, CI [3.7,4.4]), an exported protein of unknown function (2.9, CI [2.7,3.2]) and a double C2-like domain-containing protein (“DOC2”, 2.3, CI [2.1,2.5]).

**Figure 1.**
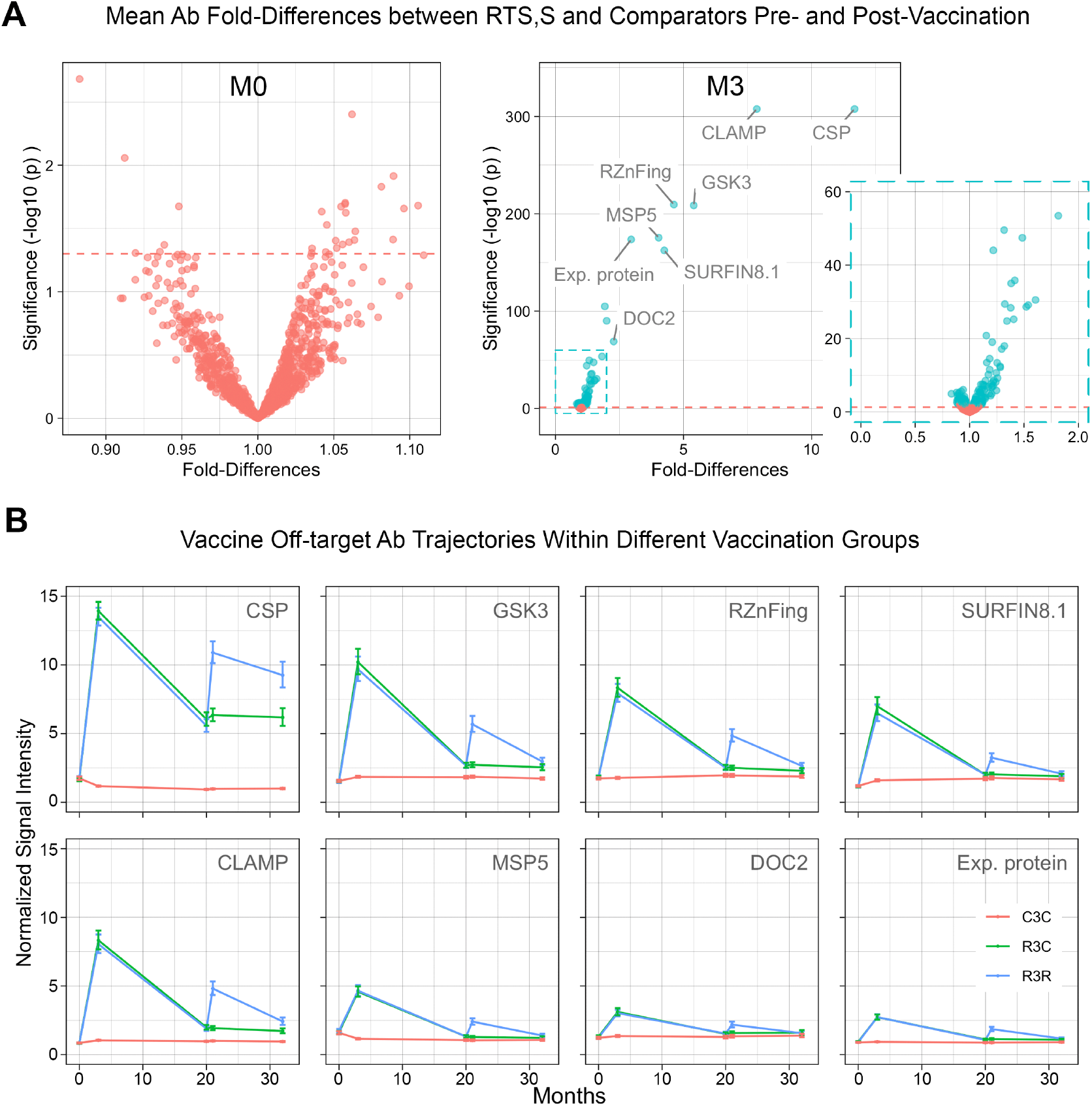
Differential antibody levels across time points. A) Volcano plots from repeated univariate regression models comparing antibody normalized signal intensity geometric means against 1,000 malarial antigens between comparator and RTS,S/AS01E groups. Effect sizes in the volcano plots are represented as fold-differences (x-axis) of RTS,S/AS01E over comparators. Green dots correspond to false discovery rate-corrected significant differences; red dots do not. LEFT: prior to vaccination (M0) when no differences were detected; RIGHT: shortly following vaccination (M3) when more than a hundred antigens were found with significant differential antibody levels. B) Longitudinal trajectories of geometric mean normalized signal intensity (i.e. log2 signal-to-noise ratio) and their 95% CI. Panels include the seven most reactive off-target antibodies at M3 in addition to CSP. Blue corresponds to RTS,S vaccination with boost at study month 20 (R3R), green without boost (R3C), red to comparator vaccination (C3C).

Further significant differences rapidly decreased in magnitude: 6 Ags had geometric mean antibody levels increased in RTS,S vaccinees of 50-100% over comparators, 14 Ags between 25% and 50%, and as many as 143 Ags with smaller differences (<25%). Of note, 70 Ags from the latter group were not increases, but decreases in the RTS,S group (green dots to the left of one in the right volcano plot, ***Figure 1A***). ***Figure 1-Supplementary Material 1*** contains details of the corresponding fold-differences between vaccination groups at M3, as well as at subsequent time points for all Ags probed in the microarray. For validation of results, a purified recombinant MSP5 protein was available and was tested in an orthologous antibody binding assay using quantitative suspension array technology (qSAT) on a subset of samples from the phase 3 trial, as well as samples from an independent phase 2b clinical trial of the RTS,S vaccine under a different formulation and with different participants. The data, which confirm the large effect size of RTS,S vaccination on MSP5 antibodies, are shown in ***Supplementary Material 2***.

### Strong vaccination-induced off-target Ab levels decline but persist over 32 months of follow-up

At M20, M21 and M32, off-target IgG levels with at least 2-fold increases at M3 (“strong” off-target Ab) remained significantly higher in the RTS,S group during follow-up time points despite a clear waning of Ab levels. ***Figure 1B*** shows their longitudinal trajectories, together with anti-CSP IgG levels for comparison. By contrast, most of the numerous Ab with small differences in levels (<2-fold) detected at M3 disappeared during follow-up or declined to an undetectable level for the sample size. Repeating the univariate screening for off-target Ab level differences at follow-up timepoints, only 7 Ags were identified at M20, 11 Ags at M21 and 17 Ags at M32 with differences passed the significance threshold, of which 5, 9 and 7 Ags, respectively, corresponded to the subgroup of strong off-target Ab at M3 (***Figure 1- Supplementary Material 1***). The few Ab differences that had not been detected at M3 but were significantly differential at later time points were always small in magnitude (<25%), mainly involved reductions in the RTS,S group and included well-known markers of malaria exposure (e.g. merozoite surface proteins “MSPs”, erythrocyte membrane proteins “EMPs”, etc).

### Booster vaccination reinforces vaccine off-target Ab increases

The RTS,S/AS01E booster dose administered 18 months after primary vaccination (M20) increased the levels of strong off-target Abs. Overall, these Ab levels mimicked the characteristic waning and increasing pattern of anti-CSP Ab after primary and booster vaccination (***Figure 1B***). Similar to anti-CSP IgG, post-booster responses to these Ags did not reach levels as high as those following primary vaccination, a characteristic of RTS,S immunogenicity (White et al., 2015, Sanchez et al., 2020). RTS,S/AS01E booster vaccination reinforced declining strong off-target Ab levels to at least 1.5-fold over comparators, with a majority above 2-fold.

Other malarial Ags also followed this pattern of increases detected following both primary and booster vaccination. In particular, 20 Abs were detected with significantly higher mean levels in the RTS,S-boosted over comparators, and nearly all had also been increased following the primary dose (***Supplementary Material 1***). The number of significant off-target Ab increases following the booster was smaller than after primary vaccination. This may be due to a combination of lower off-target Ab immunogenicity of the booster, as evident by smaller effect sizes of the induced increases, and to the decreased statistical power of the comparisons at M21 that used a smaller sample size.

### A single signature of vaccine off-target Ab responses correlated with anti-CSP Ab levels

Off-target Ab responses to vaccination mainly occurred together (cross-correlated) and displayed high levels of heterogeneity across RTS,S vaccinees. Using partial least squares discriminant analysis (“PLS-DA”) decomposition of all Ags presenting significant differences at M3, including increases and decreases, but excluding CSP from the panel, a single latent dimension of covariation with the vaccination group was identified. ***Figure 2A*** shows that beyond the 1^st^ component, cross-validated performance scores did not improve and hit a ceiling of 80-85% accuracy. Thus, vaccine-induced effects on off-target Ab levels likely did not happen independently. Vaccinees with off-target Abs strongly reacting to one Ag were very likely to also have Abs strongly reacting to other Ags in the signature.

**Figure 2.**
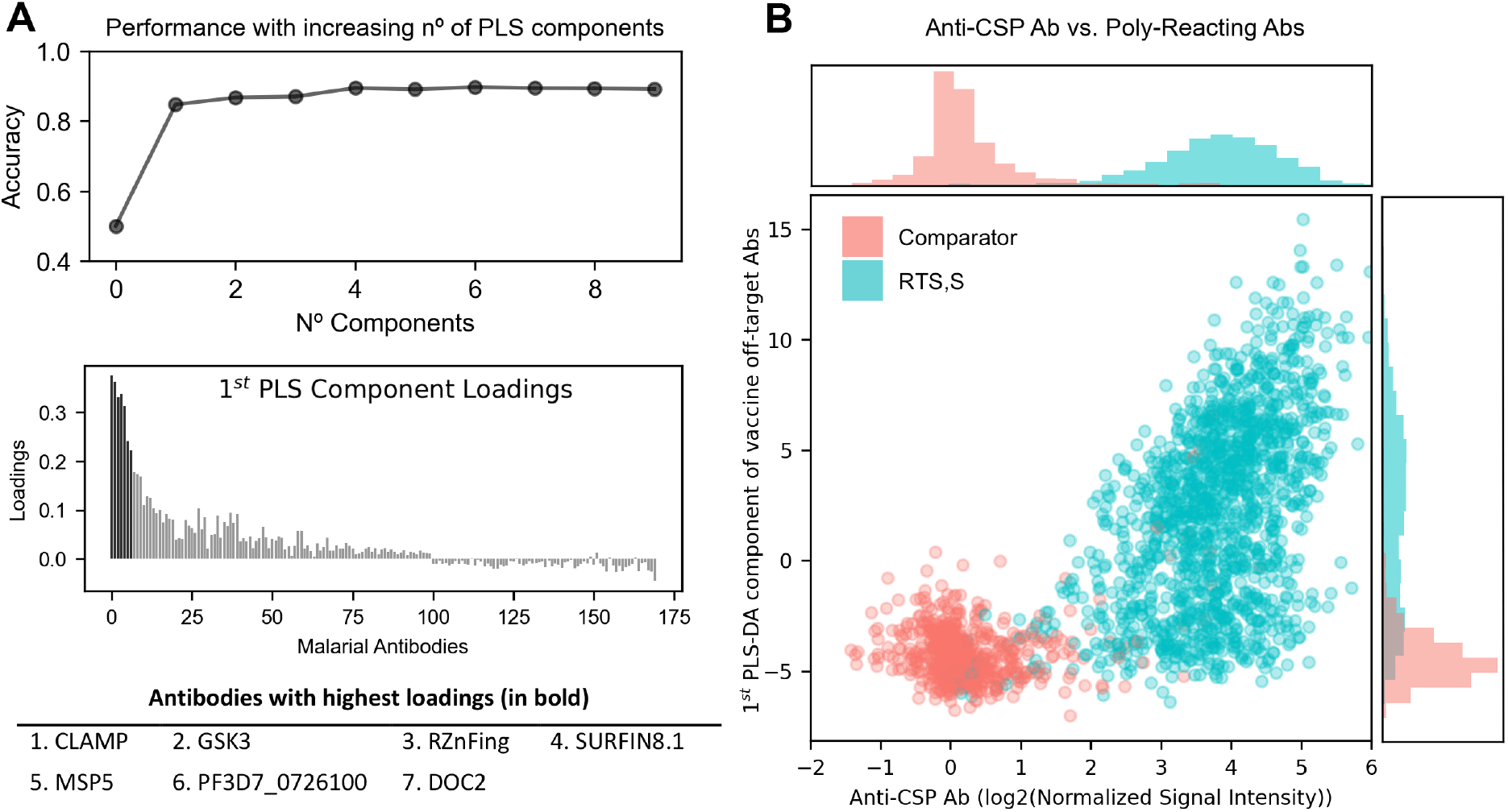
Partial Least Squares Discriminant Analysis (PLS-DA). All antigens with significant univariate differences (increases or decreases), but excluding CSP, were included as predictors in PLS-DA models with vaccination group as the outcome. LEFT, Top: cross-validated (5-fold) contribution to prediction accuracy of other components. LEFT, Bottom: loadings of antigens to the 1^st^ PLS-DA component. RIGHT: scatter plot of the 1^st^ PLS-DA component scores against CSP antibody levels. Ab: antibody.

When loadings were ranked by magnitude of fold-differences between vaccination groups, the contribution of each Ag to the signature (PLS-DA 1^st^ component loadings, ***Figure 2A***, bottom) closely resembled the univariable differences in magnitude and sign; the same pattern of strong off-target Ab increases emerged, including CLAMP, SURFIN8.1, RZnFing, etc. over a larger number of small contributions. Vaccine-induced off-target Ab changes, in addition to forming a single signature, also correlated with post-vaccination anti-CSP Ab responses. ***Figure 2B*** shows anti-CSP Ab levels against the 1^st^ PLS-DA component scores with a clear correlation within the RTS,S group (green dots, ρ=0.48, CI [0.44-0.53]), but weak within the comparator group (red dots, ρ=0.2, CI [0.12-0.26]). Thus, vaccinees with high anti-CSP Ab levels following RTS,S vaccination were more likely to present high off-target Ab increases. Using a subset of the protein array samples for which we had ELISA Ab measurements against the NANP-repeat and C-terminal (C-Term) regions of CSP separately in the vaccinees, we were able to identify that a similar correlation only existed against the NANP but not the C-Term region. Furthermore, high NANP-region Ab avidity also was associated with higher off-target Ab responses, even after adjusting for NANP Ab levels (Supplementary Material 3).

### Heterogeneity in the off-target Ab responses within RTS,S vaccinees

Within RTS,S vaccinated individuals, heterogeneity in off-target Ab responses was greater than that of anti-CSP. ***Figure 2B*** shows that the marginal histogram for off-target reactivity scores was flatter than that for anti-CSP levels. In ***Figure 3A***, comparator vaccinees (red histograms) presented Ab levels mainly centered at 0 with relatively normal unimodal distributions, some slightly skewed right. This shows that comparators were mainly seronegative for anti-CSP Ab, as well as for strong off-target Abs. In contrast, RTS,S vaccinees presented Ab levels far above background. However, whereas anti-CSP levels were homogeneously increased and nearly all subjects were seropositive, off-target Abs presented a flatter distribution extending from 0 to very high levels, indicating a highly heterogeneous response to vaccination. Surprisingly, this heterogeneity appeared in the shape of bimodal distributions, with a first mode centered near zero resembling that of comparators, and a second mode centered at high Ab levels, indicating strong seropositivity in only a subgroup of vaccinees. This characteristic was particularly notable for CLAMP, GSK3, RZnFing and SURFIN8.1.

**Figure 3.**
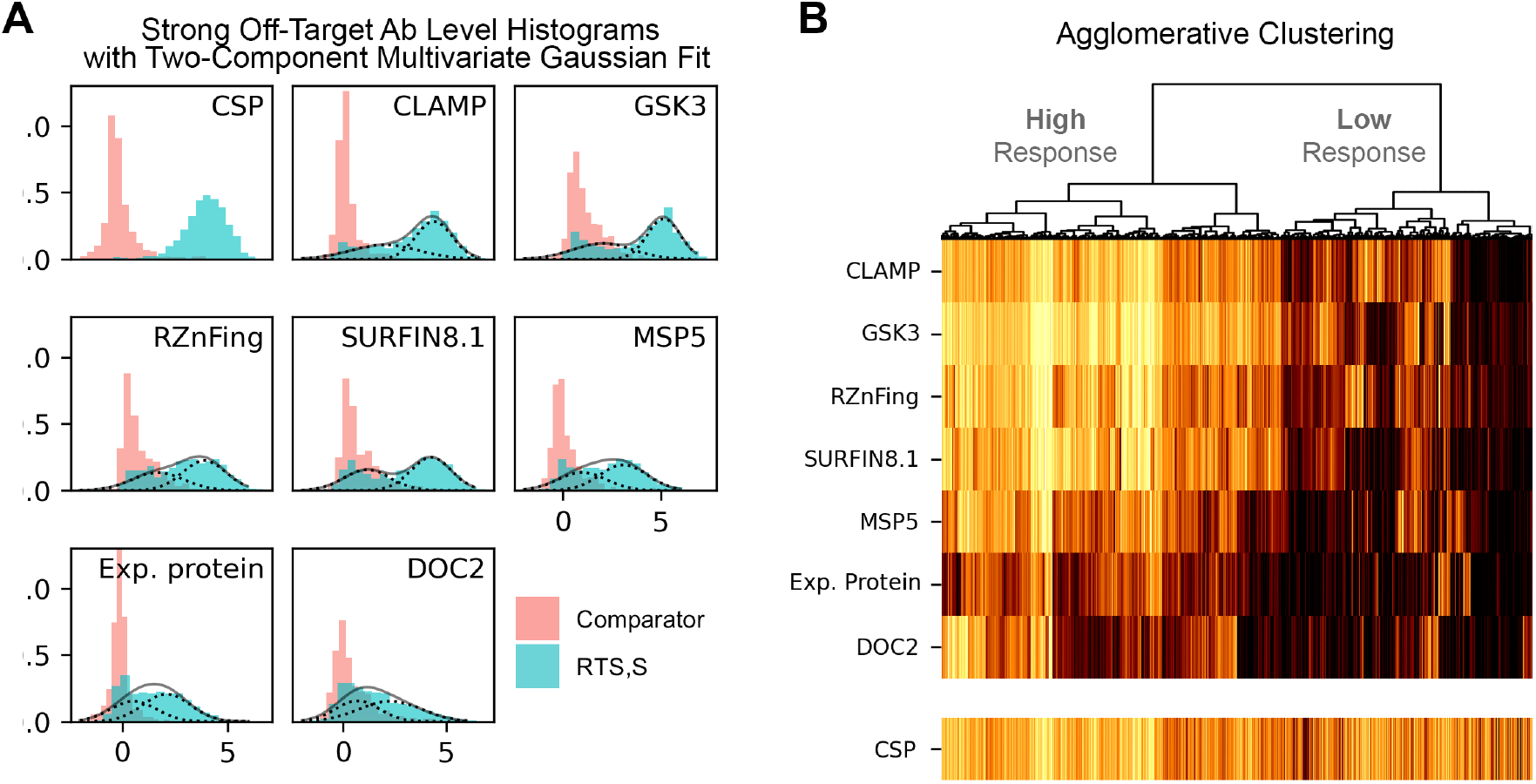
Bimodal heterogeneity in off-target antibody responses following RTS,S vaccination captured by two unsupervised classification algorithms. A) The histograms show the normalized signal intensity for anti-CSP antibodies and strong off-target antibodies at M3. A multivariate two-component gaussian mixture distribution was fitted to the data and the probability density curves for each component were overlaid (dashed lines are mixing gaussian components, solid lines are the resulting mixture). B) The heat map of normalized signal intensity for the strong off-target antibodies (excluding anti-CSP) are shown where observations (vaccinees in the columns) are ordered according to an agglomerative clustering algorithm. The dendrogram on top depicts the order of clustering merging and informs about the dissimilarity between them as branch heights are proportional to cluster distances. Anti-CSP Ab levels are plotted as a reference. Ab, antibody.

To explore if bimodality also occurred multidimensionally, i.e. seropositivity against a given off-target Ag associated with seropositivity against other off-target Ags, a two-component multivariate gaussian mixture model was fit. ***Figure 3A*** plots the predicted probability densities on top of the histograms to illustrate goodness of fit. The two-component gaussian mixture adequately captured the two sub-distributions (putative underlying subgroups) in nearly all bimodal distributions. The algorithm classified 61% of all RTS,S vaccinated individuals as high responders and provided a classification henceforth used to stratify vaccinated individuals. The binary classification was complemented with an alternative agglomerative clustering, a more agnostic algorithm that does not force the best classification to be binary. Nonetheless, ***Figure 3B*** shows that the main differences in off-target Ab responses to RTS,S vaccination was captured by the split of the top branch, confirming that a binary grouping was justified. Both classification algorithms, despite working differently, achieved 94% coincidence, a high score showing that the subgrouping of vaccinated individuals between low and high off-target Ab responders was robust.

### High off-target Ab responder group is associated with age, malaria transmission intensity and higher VE

Using the gaussian mixture binary classification, we found that children are more often “high off-target Ab responders” than infants (***Figure 4***). In high malaria transmission intensity (MTI) sites, 59.6% of children (N=184) were classified as high responders, and only 42.2% of infants (N=293) were classified as high responders, a significant difference in proportions (p=2.8e-05). The same trend was observed in low MTI sites, with more high responders in children than in infants (71.3% [N=134] and 64.1% [N=191], respectively, p=0.054). In addition, high off-target Ab responders were also more common in low than high MTI sites, as seen in the visual comparison by site and age of ***Figure 4***. However, no differences were found in proportions of high off-target Ab responders between females and males (p=0.99).

**Figure 4.**
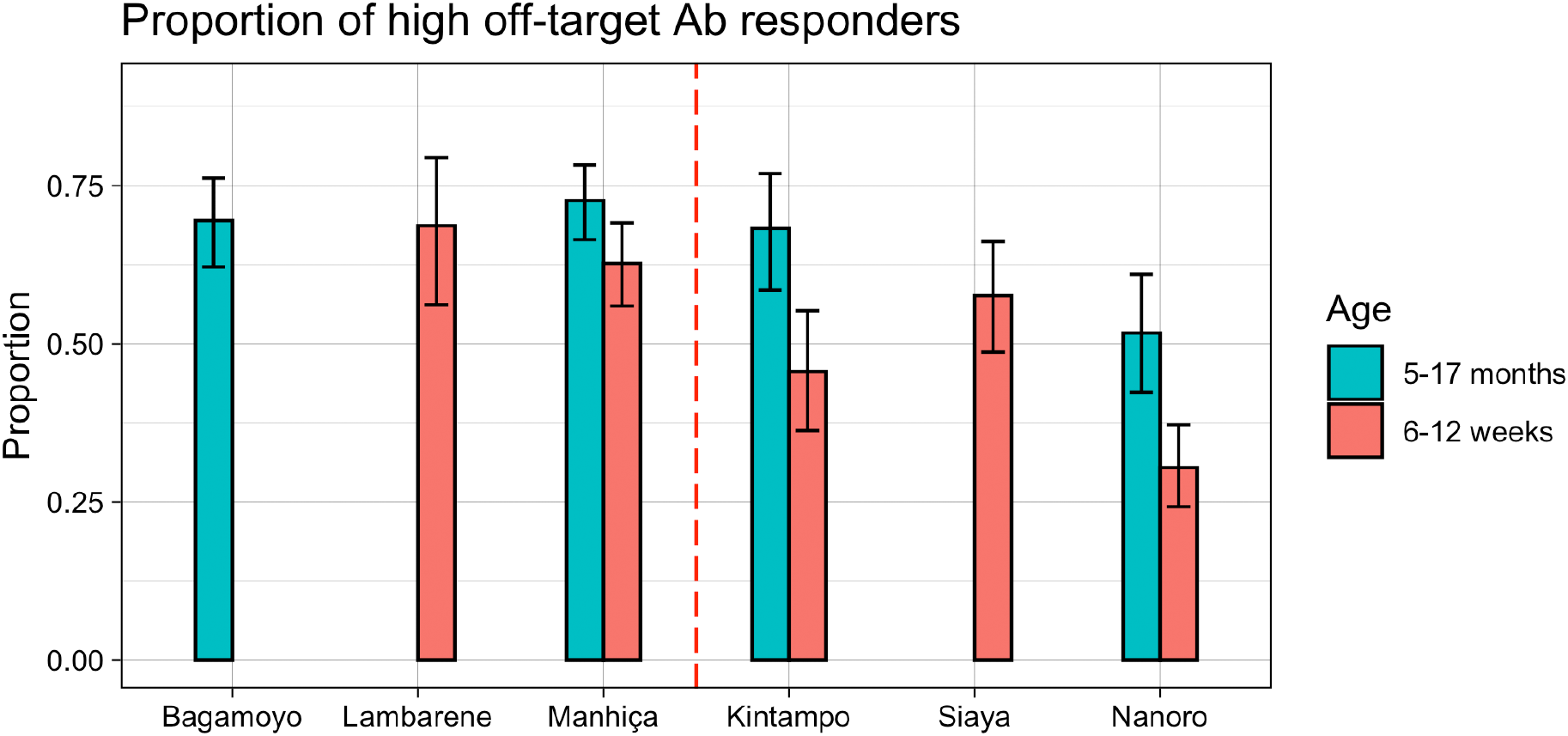
Association of vaccine off-target antibody reactivity with age group and malaria transmission intensity. Proportion of RTS,S vaccinees classified as high responders to vaccine off-target antigens is shown in the bar graph with 95% CIs for the proportions estimated as exact Clopper–Pearson binomial intervals. Missing bars are due to lack of data (i.e. samples not collected or selected) for the corresponding site.

We calculated VE over 1 year of follow-up since M3 stratified by increasing anti-CSP levels (Ab level tertiles) and off-target Ab response (low vs. high responder classification). As expected, higher anti-CSP tertiles were associated with higher VE, both in infants and children (***Figure 5***). Despite large confidence intervals in estimated VE as a consequence of low sample sizes in the RTS,S sub-groups, RTS,S-vaccinated individuals classified as high off-target Ab responders tended to have higher VE than their low responder counterparts with similar anti-CSP levels.

**Figure 5.**
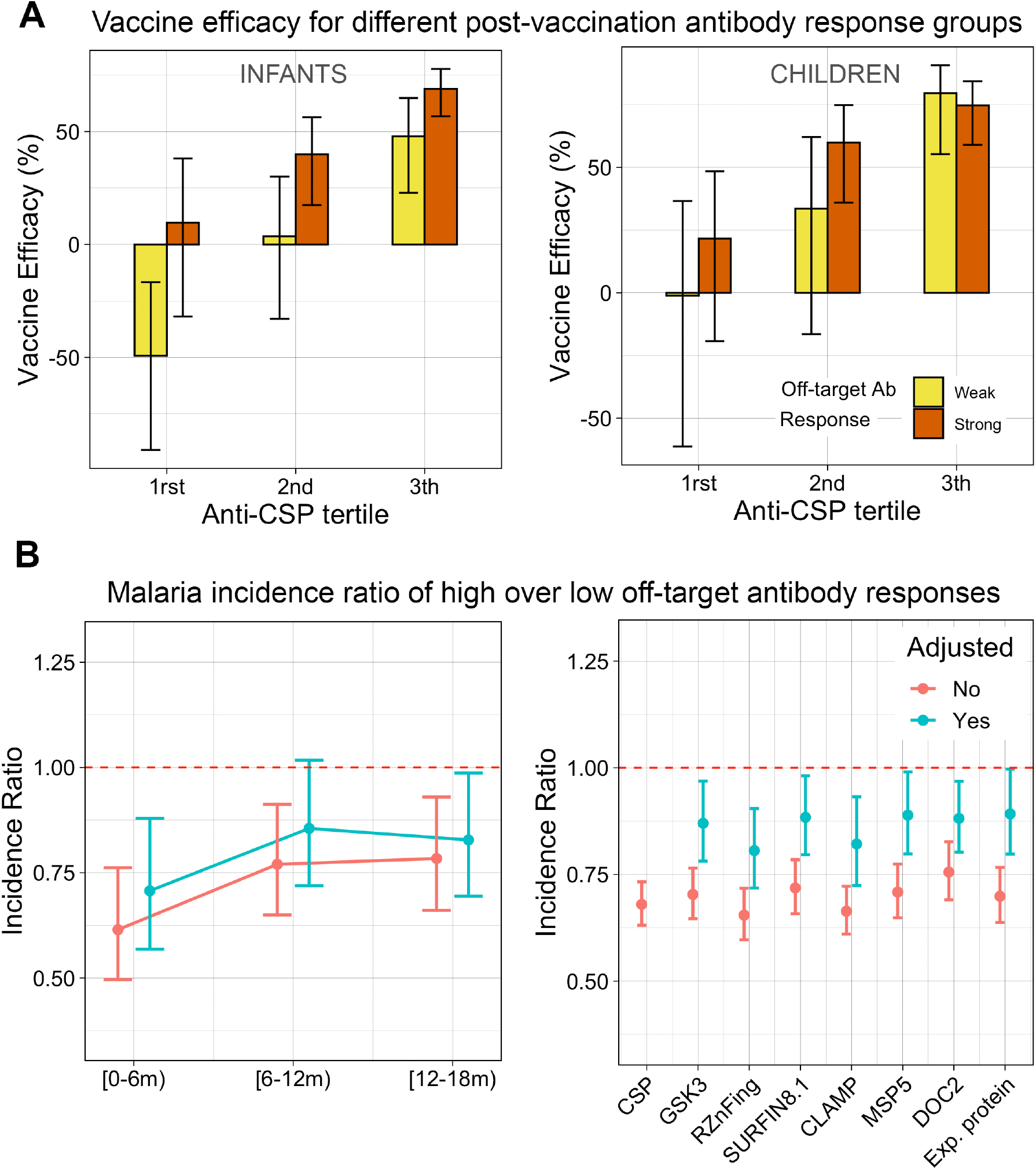
Association of the high off-target antibody responder group with vaccine efficacy. A) Vaccine Efficacy (VE) over 1 year of follow-up since M3 was repeatedly calculated for six different RTS,S sub-groups against the reference comparator group. Six post-vaccination immunological response sub-groups were defined based on anti-CSP levels (tertiles) and low vs. high vaccine off-target antibody response according to our classification. B) LEFT: clinical malaria incidence ratios were calculated within successive semesters after vaccination (M3) for high over low off-target responders. B) RIGHT: incidence ratio increase per unit change of standard deviation of normalized signal intensity for vaccine off-target antibodies with the largest increases. All plots contain estimates from models adjusted (red) and unadjusted by CSP levels (green). All models were adjusted by age group and site.

The observed trend with VE could be driven by confounding variables, mainly site or residual anti-CSP differences in the subgroups. To adjust for age, site and also anti-CSP levels, malaria incidence was modelled including these factors as control variables, and incidence ratios between RTS,S vaccinees with high over low off-target Ab responses were estimated. When adjusting for age group and site, but not for anti-CSP, malaria incidence in high off-target Ab responders was significantly lower than in their low responding counterparts for the three succeeding semesters after M3 (***Figure 5B***, left). The strongest difference in malaria incidence took place in the first semester, reaching an estimated 39% fewer cases (IR=0.61, CI [0.50-0.76], p=8.83e-06) in high off-target Ab responders; in the 2^nd^ and 3^rd^ semesters, reductions became smaller (IR=0.77, CI [0.57-0.91], p=2.53e-03, and IR=0.78, CI [0.66-0.93], p=5.20e-03, respectively). When also adjusting for anti-CSP levels at M3 (including potential non-linearities using b-spline basis functions), the association became weaker but clearly followed the same pattern, with a significant estimated malaria incidence reduction of 29% in the first semester (IR=0.71, CI [0.57-0.88], p=1.82e-03), but smaller in the following semesters (2^nd^ semester: IR=0.86, CI [0.72-1.02], p=0.08; 3^rd^ semester: IR=0.83, CI [0.69-0.99], p=0.034). This result indicates that part of the association of higher VE with vaccine off-target Ab reactivity cannot be explained by its correlation with anti-CSP levels alone.

To complement the previous analysis, instead of using the binary classification based on the joint distribution of off-target Ab responses, the association of malaria protection with strong off-target Ab levels was estimated (***Figure 5B***, right). When not adjusting for anti-CSP Ab levels, but still adjusting for age group and site, higher levels were significantly associated with protection to nearly the same degree as anti-CSP Ab levels. However, when adjusting for anti-CSP, their associations were weaker, with some no longer reaching statistical significance. No single off-target Ab clearly stood out as more predictive of protection than others.

### Sequence realignment between CSP and strongly reactive off-target Ags

A BLAST of CSP including the NANP repeat and C-Term regions against the *P. falciparum* 3D7 strain ortholog of each strongly reacting off-target Ag identified no significant sequence similarity. We repeated the analysis using HBsAg co-expressed in the vaccine instead of CSP, again obtaining no significant results. A more guided BLAST of ‘NANP’, ‘NVDP’ or combination of ‘NANPNVDP’ against full length proteins also gave no significant matches, except for CLAMP. In the C-terminal proline-rich domain of CLAMP, there is a 5 amino acid (aa) ‘NLNPN’ sequence (aa 381-385 of CLAMP) with a substitution of an alanine with leucine. To further explore short sequence alignments, we use a sliding 6 aa window (shifted one aa per window) of the ‘NANPNVDP’ sequence with MAFFT on sliding 6 aa windows of the cross-reactive Ags. The proline in the CSP repeat region was considered an irreplaceable residue for providing structural rigidity in the epitope, therefore only 6 aa sequence windows containing at least one proline were included. The top hit for CLAMP was aa 381-386, containing the ‘NLNPN’ sequence. Others had similar sequences with substitutions: for MSP5, the top sequence was an ‘NSNPNL’ at aa 111-116; for RZnFing, ‘NKNPNEN’ at aa 2072-2078; and PF3D7_0726100 had a sequence of 6 repeats each containing ‘NNNPN’ flanked by tyrosine or aspartic acid residues. For DOC2, the top sequence aligned with the ‘NVDP’ part of the CSP repeat was an ‘NNDPN’ at aa 30-34, substituting a valine for an asparagine. GSK3 and Surfin8.1 had no clear linear sequence alignments with ‘NANP’ or ‘NVDP’, however, it is possible that discontinuous sequences undetected by these methods could mimic the CSP repeat region. A limitation of this approach is our gap in knowledge of minimal epitopes for cross-reactive Abs. However, the physiochemical similarities in the top sequences of 5 of the strongly reacting off-target Ags suggests that a sequence of 5 to 6 aa may be sufficient and supports the postulate that the Ab binding to these proteins is cross-reacting IgG targeting the CSP.

## Discussion

We discovered considerable variation in Ab levels against 1,000 malarial Ags shortly after vaccination in a large subset of children from the phase 3 clinical trial of the RTS,S/AS01E malaria vaccine. Significant differences involved a large number of probed Ags (17%), of which the majority were small in effect size. To our surprise, a small subgroup of these Ags presented highly increased Ab responses in the RTS,S vaccinees, nearly as high as anti-CSP Ab (4- to 8-fold increases over comparators). Furthermore, these vaccine-induced strong increases occurred only in one part of the group of vaccinated individuals, and strong off-target Ab responses were a predictor of increased protection, beyond what anti-CSP Ab levels alone could predict. Levels of strong off-target Ab only partially waned over follow-up months. At least six were still significantly increased nearly three years after primary vaccination (M32), even in the absence of a booster dose. As with anti-CSP IgG, the booster dose served to raise mostly the same strong off-target Abs and to delay their subsequent decline.

Given the interventional nature of the study, the cause of Ab level differences between groups can be attributed to RTS,S/AS01E vaccination. However, the causal effects of vaccination on off-target Ab profiles could be of two natures: A) directly and immediately induced by vaccination, or B) indirectly and accumulating over time, due to the partial protection conferred by the vaccine, reducing exposure and thereby differentially acquired immunity. The temporal proximity of sampling after vaccination (M3 or M21) indicates that the majority of vaccine off-target Ab differences were directly induced by vaccination. In addition, at least two different biological mechanisms are possible for a direct effect of vaccination: 1) the large number of short-lived, small differences (<2-fold), including Ab increases and decreases, may result from a systemic perturbation of the immune system following vaccination, probably due to the immunostimulatory properties of the adjuvant, and the subgroup of strong off-target Ab increases, comparable in magnitude to anti-CSP levels, may arise from epitope similarities between the RTS,S protein target and certain off-target Ags (i.e. cross-reactivity). Previous studies have shown that cross-reactivity between epitopes located in different malaria proteins is possible (Hope et al., 1984; Anders et al., 1986). The fact that the intensity of the vaccine off-target Ab response followed a single signature and strongly correlated with post-vaccination anti-CSP IgG levels, specifically with the NANP region, supports this hypothesis. Sequence realignment analysis identified NANP-like stretches containing N and P repeats in some of these strongly reacting off-target Ags, in agreement with evidence that cross-reacting epitopes usually occur between immunodominant low-complexity repeat structures rich in asparagine and glutamate (Ardeshir et al., 1990; Hou et al., 2020). Their abundance in the malaria proteome is hypothesized to play a role in the evasion of the host’s immune response (Feng et al., 2006; Renia et al., 2016)

Discarding the possibility of differential exposure indirectly influencing off-target Ab binding can be justified due to the assumption that reduced exposure in RTS,S vaccinees is expected to cause reduced anti-malarial Abs, not increases. The hypothesis that increased subpatent parasite exposure due to partial protection afforded by RTS,S/AS01E vaccination would cause the increase in off-target Abs is also unlikely, in that sufficient exposure is unlikely to have occurred during primary vaccination or directly following the booster to elicit these responses, particularly in low MTI settings. Additionally, the off-target Ab responses decayed over follow-up, rather than accumulate. Off-target reactive Ags did not particularly include typical markers of malaria exposure, such as apical merozoite antigen 1 (AMA1) or merozoite surface proteins (MSPs but excluding MSP5), which were decreased in RTS,S vaccinees as expected. Nearly all comparator vaccinees were seronegative for the strong reacting off-target Ags (their Ab levels were close to background). Taken together, the off-target Abs observed following RTS,S/AS01E immunization were most likely direct effects of vaccination.

Vaccine off-target reactivity also presents intriguing characteristics. Off-target Ab increases were cross-correlated, defining an “off-target Ab response signature” with heterogeneous responses for different vaccinated individuals. Because the intensity of this response also correlated with post-vaccination anti-CSP levels, a parsimonious explanation could be that anti-CSP Abs and strongly reacting off-target Abs are the same, and the phenomenon would be a typical example of Ab cross-reactivity. However, this does not explain why there exists a substantial group of vaccinated individuals (around 40%) with very low vaccine off-target Ab levels, nearly as low as comparator vaccinees, whilst their anti-CSP Ab levels can be high or even very high. We captured this binary subgrouping of high vs. low off-target Ab responders with multiple clustering techniques, although the pattern is most easily seen in the bimodal distributions of post-vaccination off-target Ab levels in the RTS,S group. When a categorical grouping neatly emerges from data, it is often indicative of an underlying categorical determinant. However, we cannot rule out that bimodality may alternatively arise from a measurement limitation in the protein array technology. Yet, whether the heterogeneity in off-target Ab reactivity in vaccinated individuals is amenable to a binary subgrouping or, in reality, is not multimodal and simply spans a continuum, has little implications in the relevance of these results.

Vaccinated individuals presenting high off-target Ab responses had an estimated lower clinical malaria incidence. This association with protection was partially maintained as we adjusted, for age, site and post-vaccination anti-CSP levels. In other words, vaccinated individuals with a high off-target Ab response to RTS,S/AS01E vaccination were more protected from infection than their vaccinated counterparts with similar age, site and anti-CSP Ab levels. The result that high off-target Ab responses is associated with lower estimates of malaria incidence suggests that this immune signature may confer additional benefits, aside from being a proxy of high anti-CSP Ab levels, the best serological marker of VE thus far (White et al., 2015). One explanation is that vaccine off-target Abs binding to malarial Ags, possibly just anti-CSP IgG that are cross-reacting with them, are indeed playing a biological role in preventing symptomatic infections. CLAMP (in micronemes), SURFIN8.1 (in cell membrane), MSP5 (merozoite surface), DOC2 (host cell plasma membrane), GSK3 (at Maurer’s cleft), are all cell surface proteins expressed at different stages of the *P. falciparum* parasite life cycle. Mounting an Ab-led immune response against them, assuming some must be sufficiently exposed at a certain stage of the parasite’s life cycle, could potentially be beneficial and reduce parasitemia. However, our observational result is also compatible with another scenario in which vaccine off-target Abs do not play any mechanistic role in protection and would simply be a proxy of the quality of the CSP response or other underlying protecting factors yet to be discovered. One of such factor could be Ab maturation, since we found an association of high NANP Ab avidity with high Ab off-target responses after adjusting for anti-NANP Ab concentrations in a subset of samples with additional avidity measurements, (Supplementary Material 3).

The present study has three main implications for future research. First, it forewarns researchers investigating post-vaccination naturally acquired immunity in populations vaccinated with RTS,S or other CSP-based vaccines of misinterpreting vaccine-induced off-target Ab changes. These studies usually focus on mid-to long-term immunological estimates and will likely encounter the long-lasting effects of vaccine-induced off-target Ab increases, which should not be mistakenly interpreted as markers of differential exposure in vaccinated populations. Second, having established that RTS,S-induced Abs bind not only to CSP but to other unrelated malarial antigens, further research should investigate whether some of the less characterized Ags are sufficiently exposed within the parasite and crucial to its lifecycle for such off-target Abs to be beneficial to the human host. Third, further research is needed to determine whether off-target Ab reactivity is a common phenomenon also present after vaccination with other subunit vaccines and whether it is indeed caused by cross-reactivity, as we suspect at least for the strong off-target Abs. If the latter was to be the case, then this discovery would have far-reaching implications beyond RTS,S/AS01E vaccination and malaria.

Cross-reactivity often occurs between strains of the same pathogen, close but different species, and even phylogenetically separated ones (heterologous or between-pathogen cross-reactivity) (Vojtek et al., 2019). Edward Jenner’s historical discovery of a vaccine against smallpox using inocula from cowpox was made possible, unknowingly, thanks to it. But, to our knowledge, no previous research has aimed to investigate the possibility of within-pathogen cross-reactivity. By this term, we mean the capacity of a monoclonal Ab that has matured to bind to a cognate Ag from a given pathogen to additionally bind, perhaps with lower affinity, to other non-cognate Ags from the same pathogen. Such a phenomenon may not be uncommon in nature (Dobaño et al., 2021; Requena et al., 2017) but may have remained unnoticed due to lack of interest or technological limitations allowing broad antigen screening. However, the paradigm of the one-to-one ‘lock and key’ antigen–antibody interaction is shifting towards a more nuanced ‘several-to-several’ model. Some degree of cross-reactivity is finally acknowledged as 1) not uncommon if Abs are screened against a sufficiently large number of Ags, 2) potentially beneficial for the host and perhaps even favoured by Ab clonal selection itself (Guthmiller et al., 2020; Agrawal, 2019).

In this study, we have shown that high throughput Ab screening in the course of vaccine trials can reveal broad vaccine-induced off-target Ab alterations, with some of these off-target Abs reaching impressive increases that are compatible with strong within-pathogen cross-reactivity. Importantly, the presence of strong off-target Abs was also a correlate of increased protection. Their corresponding antigens could potentially be candidates of multivalent next-generation RTS,S formulations as additive (or synergistic) responses with CSP may occur.

## Materials and Methods

### Study Design and Data

This study was carried out in a subset of the phase 3 randomized clinical trial MAL055 (NCT00866619) and the MAL067 immunology study including infants (age 6-12 weeks at enrolment) and children (age 5-17 months at enrolment) from 6 different African sites. The clinical trial tested safety, immunogenicity and VE (RTS,S and Partnership, 2015; White et al., 2015). Participants received 3 doses of either the RTS,S/AS01E vaccine or a comparator vaccine (the meningococcal C conjugate in infants or rabies vaccines in children) at study months (M) 0, 1, and 2 for primary vaccination, and a booster dose at month 20 (M20). In the MAL067 study, blood samples were collected prior to the start of primary vaccination (M0); three months after (M3, one month after third dose); before the booster dose (M20); one month after it (M21), and at the end of follow-up (M32). Further details of sample size and demographic characteristics of the subsamples analysed at each timepoint are reported in ***Table 1***.

**Table 1.** Sample size and comparison of descriptive statistics between the different vaccination groups at each time point for samples probed on microarrays.

|  | M0 |  | M3 |  | M20 |  | M21 |  |  | M32 |  |  |
| --- | --- | --- | --- | --- | --- | --- | --- | --- | --- | --- | --- | --- |
|  | C | R | C | R | C | R | C3C | R3C | R3R | C3C | R3C | R3R |
| **Sample Size** | 512 | 1048 | 693 | 1376 | 343 | 720 | 377 | 428 | 374 | 368 | 230 | 226 |
| **Sex (% Females)** | 55.7 | 50.0 | 54.7 | 50.9 | 53.4 | 48.6 | 54.1 | 49.5 | 45.2 | 52.7 | 50.4 | 46.5 |
| **Age (% Infants)** | 57.0 | 58.2 | 52.8 | 53.8 | 50.7 | 52.9 | 40.3 | 41.4 | 43.3 | 42.9 | 41.3 | 40.7 |
| **Site (% Low MTI)** | 54.9 | 52.5 | 53.1 | 50.7 | 33.5 | 33.1 | 42.2 | 42.5 | 37.7 | 47.8 | 63.9 | 61.1 |
| * **Manhiça** | 36.7 | 34.6 | 35.1 | 33.0 | 33.5 | 33.1 | 20.7 | 20.3 | 17.9 | 28.3 | 43.9 | 40.7 |
| * **Lambaréné** | 1.2 | 1.2 | 5.2 | 4.9 | 0.0 | 0.0 | 0.0 | 0.0 | 0.0 | 0.0 | 0.0 | 0.0 |
| * **Bagamoyo** | 17.0 | 16.7 | 12.8 | 12.9 | 0.0 | 0.0 | 21.5 | 22.2 | 19.8 | 19.6 | 20.0 | 20.4 |
| **Site (% High MTI)** | 45.1 | 47.5 | 46.9 | 49.3 | 66.5 | 66.9 | 57.8 | 57.5 | 62.3 | 52.2 | 36.1 | 38.9 |
| * **Kintampo** | 14.3 | 13.7 | 24.4 | 23.6 | 43.2 | 40.8 | 38.7 | 35.5 | 38.8 | 34.8 | 17.4 | 19.9 |
| * **Siaya** | 12.7 | 12.6 | 9.5 | 9.6 | 0.0 | 0.0 | 0.0 | 0.0 | 0.0 | 0.0 | 0.0 | 0.0 |
| * **Nanoro** | 14.3 | 13.7 | 24.4 | 23.6 | 43.2 | 40.8 | 38.7 | 35.5 | 38.8 | 34.8 | 17.4 | 19.9 |
| **Lost to follow-up (%)** |  |  |  |  |  |  |  |  |  |  |  |  |
| * **6 months** |  |  | 3.3 | 1.7 |  |  |  |  |  |  |  |  |
| * **12 months** |  |  | 6.3 | 3.9 |  |  |  |  |  |  |  |  |
| * **18 months** |  |  | 8.8 | 6.6 |  |  |  |  |  |  |  |  |

Sex, age group and site were used as covariates either of interest or for adjustment purposes. Sites were classified as high or low malaria transmission intensity: Bagamoyo, Lambaréné and Manhiça (representative of moderate/low MTI) had average annual malaria incidences in the non-RTS,S vaccinated group after M3 of 0.2, 0.2, 0.1 respectively; Kintampo, Nanoro and Siaya (high MTI) of 2.3, 3.0 and 3.6. To model malaria incidence, we used the trial secondary clinical malaria case definition: illness in a child brought to a study facility with a measured temperature of 37.5°C or more, or reported fever within the past 24 h, and *P. falciparum* asexual parasitaemia at any density. No new episodes were registered during 14 days after an episode which met the case definition under evaluation to account for the short-term chemoprophylactic effect of antimalarial treatment. Furthermore, to correct for time at risk, the same 14 days were subtracted from the corresponding individual’s follow-up time.

The multicenter study protocol was approved by the Comitè Ètic d’Investigació Clínica (CEIC, Hospital Clínic Research Ethics Commitee, ref 2008/4622), Barcelona, Spain, and the Research Ethics Committee (REC) for PATH, USA. Written informed consent was obtained from parents or guardians before the start of the work.

### Antigen Microarray Data

The design of the protein microarray and procedures used in this study have been described elsewhere (Kessler et al., 2018). Briefly, a partial proteome microarray with 1,000 *P. falciparum* protein features (Pf1000, 3rd generation) was developed at Antigen Discovery, Inc. (ADI, Irvine, CA, USA). The 1,000 full-length or partial *P. falciparum* proteins represent 762 genes from *P. falciparum* reference strain 3D7, including 61 PfEMP1s, vaccine candidate proteins, and 176 conserved *Plasmodium* proteins of unknown function.

Proteins were expressed using an *in vitro* transcription and translation (IVTT) system, the *Escherichia coli* cell-free Rapid Translation System (RTS) kit (5 Prime, Gaithersburg, MD, USA). A library of partial or complete open reading frames (ORFs) cloned into a T7 expression vector pXI has been established at ADI. This library was created through an *in vivo* recombination cloning process with PCR-amplified ORFs, and a complementary linearized expressed vector transformed into chemically competent *E. coli* was amplified by PCR and cloned into pXI vector using a high-throughput PCR recombination cloning method described elsewhere (Davies et al., 2005). Each expressed protein includes a 5’ polyhistidine (HIS) epitope and 3’ hemagglutinin (HA) epitope. After expressing the proteins according to manufacturer instructions, translated proteins were printed onto nitrocellulose-coated glass AVID slides (Grace Bio-Labs, Inc., Bend, OR, USA) using an Omni Grid Accent robotic microarray printer (Digilabs, Inc., Marlborough, MA, USA). Each slide contained eight nitrocellulose “pads” for which the full array was printed in replicate, allowing eight samples to be probed per slide. Microarray chip printing and protein expression were quality checked by probing random slides with anti-HIS and anti-HA monoclonal Abs with fluorescent labelling.

Prior to sample application, a probing plan was developed to balance serum/plasma samples across microarray slides by the following variables: sample collection time point, clinical trial site, age group, sex and case-control status during the first 12 months of follow-up post-vaccination. Paired samples from a single trial participant were probed onto a single chip (8 samples per chip; up to 5 paired samples per chip) or subsequently printed chips to minimize variation for time point comparisons.

Samples were probed on the Pf1000 microarrays at 1:100 with a 3mg/mL DH5α *E. coli* lysate (GenScript) overnight at 4°C. Secondary anti-IgG Ab (biotin-SP-conjugated donkey anti-human IgG; Jackson Immuno) was applied the following day at 1:1,000 (respectively in 2% *E. coli* lysate; GenScript) for 1 h at room temperature. Streptavidin Sensilight-P6824 (Columbia Biosciences) was applied to each slide at 10µg/mL (2% *E. coli* lysate; GenScript) for 1 h, covered (no light) at room temperature. Washes were performed with TTBS (Tween Tris-buffered saline) 3x before/after all incubations. Prior to overnight drying in a desiccator, all slides were washed with TBS (Tris-buffered saline) 3x and spun dry by centrifugation at 1,000 x g for 4 min. Air dried chips were scanned on a GenePix 4300A High-Resolution Microarray Scanner (Molecular Devices, Sunnyvale, CA, USA), and spot and background intensities were measured using an annotated grid file (.GAL). Data were exported in Microsoft Excel.

Raw spot and local background fluorescence intensities, spot annotations and sample phenotypes were imported and merged in the R statistical environment (www.r-project.org), where all subsequent procedures were performed. Foreground spot intensities were adjusted by local background by subtraction, and negative values were converted to 1. Next, all foreground values were transformed using the base 2 logarithm. The dataset was normalized to remove systematic effects by subtracting the median signal intensity of the IVTT controls for each sample. Since the IVTT control spots carry the chip, sample and batch-level systematic effects, but also antibody background reactivity to the IVTT system, this procedure normalizes the data and provides a measure of the specific antibody binding relative to the non-specific antibody binding to the IVTT controls (a.k.a. background). Thus, the normalized signal intensity is the log2-transformed signal-to-noise ratio, whereby a value of 0 represents antibody levels equivalent to the background, a value of 1 represents antibody levels twice that of the background, and each subsequent unit increase represents a doubling of antibody levels.

### Data Analysis

#### Univariate Ab differences between vaccination group

For each time point and Ab response to the microarray antigens, we fitted a linear model with vaccination group as our variable of interest and age group, site and sex as control variables to account for their slight imbalances between vaccination groups (***Table 1***). Our outcome variable was the *normalized signal intensity* of each probed antigen in the microarray. Therefore, the estimated linear coefficient encoding the vaccination group difference, when exponentiated, could be interpreted as point estimates of geometrical mean fold differences in Ab levels of one vaccination group over the other, holding the control variables fixed. Regression coefficient standard deviations were used to generate 95% CI. We only reported differences that were significant at a false discovery rate of 5% following the Benjamini-Hochberg method (Benjamini and Hochberg, 1995).

#### Partial Least Squares Discriminant Analysis (PLS-DA) to capture post-vaccination Ab latent potential signatures

We conducted a partial least square decomposition of all microarray responses to malarial antigens to find latent signatures that maximally correlated with the vaccination group. We removed anti-CSP Ab levels from the panel to explore covariation with levels beyond the RTS,S target Ag. We used the PLS1 algorithm with 500 maximum number of iterations of the NIPALS inner loop as is implemented by Scikit-learn (version 0.20). To study the number of relevant latent dimensions of covariation with the vaccination group, we cross-validated (5-fold) the prediction accuracy of our PLS-DA with an increasing number of components.

#### Unsupervised classification of vaccinated individuals according to their post-vaccination Ab profiles

We used two different unsupervised algorithms to classify RTS,S/AS01E vaccinees into two subgroups according to their increase in off-target Ab levels following vaccination, again excluding anti-CSP from the panel:

1. Multivariate two-component gaussian mixture. Motivated by the conspicuous bimodal distributions of the log-transformed Ab MFI levels, we used a two-component multivariate gaussian fit, with estimated full covariance (no constraints) for each component, to model an empirical probabilistic distribution based on two underlying classes with different mean, covariance and assumed normal distribution. This empirical distribution allowed us to classify our observations based on their probability to belong to two subgroups of vaccinees, as well as to classify unseen new data.
2. Hierarchical agglomerative clustering. We repeated the same analysis using a more agnostic classification algorithm that does not require an a priori choice of the number of resulting classes. We ran a hierarchical clustering that iteratively merged data observations pair-wise, that is RTS,S vaccinees according to their post-vaccination off-target Ab levels, into increasingly large clusters, ordering the merges using the Ward variance minimization algorithm (Müllner, 2011). Thus, we obtained successive partitions or clusters, hierarchically ordered from small to large and outputted a measure of distance between merging clusters at each level. The method, combined with an adequate visual representation such as dendrograms and sorted heatmaps, can be very helpful to explore multivariate relations, not necessarily linear, and visualize the emergence of types and subtypes.

For both unsupervised classification algorithms, we used their Scikit-Learn implementation (Scikit-learn, 2021). Further details about algorithm initialization parameters can be viewed in the documented analysis code.

### Association of clinical malaria incidence with vaccine off-target Ab signatures

We modelled clinical malaria incidence (number of infections and re-infections over a follow-up time) with a negative binomial regression, a probabilistic model where the number of infections and re-infections is assumed to follow a negative binomial distribution with the logarithm of the average incidence conditional on a weighted sum of predictors. We added the logarithm of the follow-up time as an offset variable to account for different times at risk between individuals. Having used a log link function, the exponentiated regression coefficients automatically become incidence ratios (IR) of a given level in a categorical predictor over the reference level. For continuous predictors, estimated IRs refer to the comparison of malaria incidence within two populations with a unit change difference in the continuous predictor. Whenever we estimated IR for continuous variables, we standardized the predictor, i.e., unit changes became unit standard deviation increases (or decreases). We also used negative binomial regression to estimate VE for different sub-groups of RTS,S vaccinees classified according to their vaccine off-target Ab levels with the usual formula: VE = 1 - IR, where IR, in this case, specifically involves the ratio of malaria incidence within the RTS,S sub-group of interest over that of the comparator group. We used the glm.nb function from the MASS R package (Ripley et al., 2018) to fit negative binomial regressions to our data.

### Data Characteristics

RTS,S and comparator vaccinees in our study subsample were similar with regards to pre-treatment demographic and epidemiological characteristics such as age group, sex and site (***Table 1***). In particular, imbalances were very small (<1%) for age group, but slightly greater for site and sex (<5%) as regards to all time points except for M32 when a considerable difference in proportion of sites existed (15%). To prevent potential biases, we adjusted for these variables in all analyses comparing vaccination groups. Malaria incidence was calculated using all participants who had M3 samples. Follow-up intervals always started at M3 (blood sampling, performed approximately one month following the end of primary vaccination) and ended 6, 12 or 18 months afterwards depending on analysis. Participants who were lost to follow-up were less than 5% in either vaccination group (***Table 1***). The analysis involving malaria protection or VE was restricted to participants who were not lost to follow-up (complete case analysis). However, the analysis was repeated including lost-to-follow-up individuals, after adjusting for their shorter time at risk in the off-set of the negative binomial, to show robustness against the small number of dropouts (***Supplementary Material 4***)

## Data Availability

All data produced in the present work are contained in the manuscript

## Acknowledgements

We are grateful to the volunteers and their families, the clinical, field and lab teams at the research institutions, and the MAL067 Vaccine Immunology Consortium investigators and Working Groups. We thank the team at Antigen Discovery, Inc. that performed the microarray experiments, including Jozelyn Pablo, Chris Hung and Andy Teng for microarray fabrication, to Adam Shandling and Jonathon Truong for performing the microarray probing experiments. We thank Núria Díez-Padrisa, Rebeca Santano, Ruth Aguilar, Matthew McCall, Inocencia Cuamba, Ross Coppel, Miquel Vázquez, Evelina Angov and Peter Crompton for their contributions to the study. We thank GlaxoSmithKline Biologicals SA for their support, in 2008-2009, for developing the study plan and for transferring custodiancy of the MAL067 samples to ISGlobal. GlaxoSmithKline Biologicals SA was provided the opportunity to review a preliminary version of this manuscript for factual accuracy, but the authors are solely responsible for final content and interpretation.

The study was supported by funds from NIH-NIAID (USA, R01AI095789), PATH-Malaria Vaccine Initiative, and the Ministerio de Economía y Competitividad (Instituto de Salud Carlos III, PI11/00423 and PI17/02044) cofounded by FEDER funds/European Regional Development Fund (ERDF). This research is part of the ISGlobal Program on the Molecular Mechanisms of Malaria which is partially supported by the Fundación Ramón Areces. We acknowledge support from the Spanish Ministry of Science and Innovation through the “Centro de Excelencia Severo Ochoa 2019-2023” Program (CEX2018-000806-S), and support from the Generalitat de Catalunya through the CERCA Program.

## Supplementary Material 2. Merozoite Surface Protein 5 (MSP5) antibody (Ab) increases following RTS,S vaccination measured with different immunoassays and in another clinical trial

### 1. Presentation of replication datasets

We investigated MSP5 Ab increases one month following RTS,S primary vaccination in two other datasets for which we also had MSP5 Ab measurements:

1. MSP5 IgG responses from 195 young children and infants from a nested immunological study of the phase 3 clinical trial of the RTS,S/AS01E vaccine (MAL055 NCT00866619) using quantitative suspension array technology (qSAT) (ref: Dobaño BMED).
2. MSP5 IgGs responses from 813 children age 1-4 yrs old from a nested immunological study of a phase 2b clinical trial of the RTS,S/AS02 vaccine using qSAT (Jairoce C, in preparation)

Dataset 1) is a subsample of the same phase 3 vaccine trial from which the microarray samples were obtained in the main manuscript. The overlap between the two studies is only partial: out of 195 qSAT samples, 139 children were also included in the microarray substudy. Therefore dataset 1) can be regarded as a replication in a subset of nearly the same data using a different measurement technology.

By contrast, 2) is a completely independent dataset, with different participants, of a different age range, and a different formulation of the RTS,S vaccine in terms of the adjuvant.

Other off-target antigens that were found increased in the microarray were not included in the panels from the above studies and could not be replicated.

### 2. Three MSP5 protein constructs probed with the antigen microarray

In the 1000 *P. falciparum* microarray used for the manuscript, we assayed three different constructs from the full MSP5 protein. Below their a.a. sequences:

#### MSP5 (i), PF3D7_0206900.1.1o2

MNILCILSYIYFFVIFYSLNLNNKNENFLVVRRLMNDEKGEGGFTSKNKENGNNNRNNENELKEEGSLPTKMNEKNSNSSDKQPNDISHDESKSNSNNSQNIQKEPEEKENSNPNLDSSENSSESATRSVDISEHNSNNPETKEENGEEPLDLEINENAEIGQEPPNRLHF

#### MSP5 (ii), PF3D7_0206900.1.e1

MNILCILSYIYFFVIFYSLNLNNKNENFLVVRRLMNDEKGEGGFTSKNKENGNNNRNNENELKEEGSLPTKMNEKNSNSSDKQPNDISHDESKSNSNNSQNIQKEPEEKENSNPNLDSSENSSESATRSVDISEHNSNNPETKEENGEEPLDLEINENAEIGQEPPNRLHFD

#### MSP5 (iii), PF3D7_0206900.1.2o2

NVDDEVPHYSALRYNKVEKNVTDEMLLYNMMSDQNRKSCAINNGGCSDDQICININNIGVKCICKDGYLLGTKCIILNSYSCHPFFSILIYITLFLLLFV

As reported in the manuscript and ***Supplementary File 1***, MSP5 (i) was strongly increased following vaccination (M3), 4.0, 95% CI [3.7, 4.4]. MSP5 (ii) was also increased but less (1.99

CI [1.87, 2.13]). Finally, MSP5 (iii) was not increased significantly (1.01 CI [0.98, 1.04]).

In the replication datasets, we measured IgG responses using another MSP5 construct, that is the same for both studies but different from those in the microarray. Below we reference its a.a. sequence, highlighting in yellow the overlap with the microarray MSP5(i), in red with MSP5(ii) and underscored with MSP(iii):

MNILCILSYIYFFVIFYSLNLNNKNENFLVVRRLMNDEKGEGGFTSKNKENGNNNRNNENELKEEGSLPTKMNEKNSNSSDKQPNDISHDESKSNSNNSQNIQKEPEEKENSNPN LDSSENSSESATRSVDISEHNSNNPETKEENGEEPLDLEINENAEIGQEPPNRLHFD<u>NV DDEVPHYSALRYNKVEKNVTDEMLLYNMMSDQNRKSCAINNGGCSDDQICININNIGVKCICKDGYLLGTKCIILNSYSCHPFFSILIYITLFLLLFV</u>

The difference between MSP5 (i) and MSP5 (ii) of a single additional aspartic acid (“D”) is notable because the additional amino acid is separated from the prior string by an intron in the current annotation on PlasmoDB.org. Thus, since genomic DNA was used for cloning, lower cloning efficiency in MSP5 (ii) may account for the difference in signal strength.

### 3. MSP5 IgG increases from a nested immunological study of the same phase 3 RTS,S/AS01E vaccine trial using qSAT

We measured IgG responses, assayed using qSAT applying the xMAP™ technology (Luminex Corp., Texas), from 195 young children vaccinated with either RTS,S/AS01E (n=129) or a comparator vaccine (n=66), one month after the last dose of the primary vaccination (M3). Further details on sample characteristics and data acquisition can be obtained from published results (Dobaño et al., 2019).

For this study, we estimated a four times larger geometric mean of MSP5 Ab levels of RTS,S over comparator vaccinees one month following primary (fold-increase of 4.0, 95% CI [2.4, 6.6]). Of note, the point estimate fold increase is here very similar to the one we estimated for the protein construct MSP5 (i) from the antigen microarray.

Out of 195 qSAT samples, 139 children were also included in the microarray substudy, allowing us to pair both measurements and investigate their agreement. Figure 1 shows that the greatest correlation of the qSAT MSP5 measurements is with the MSP5 (i) protein construct of the microarray. This result supports that the putative cross-reacting epitope should lie in their overlapping sequences (highlighted in yellow).

**Figure 1.**
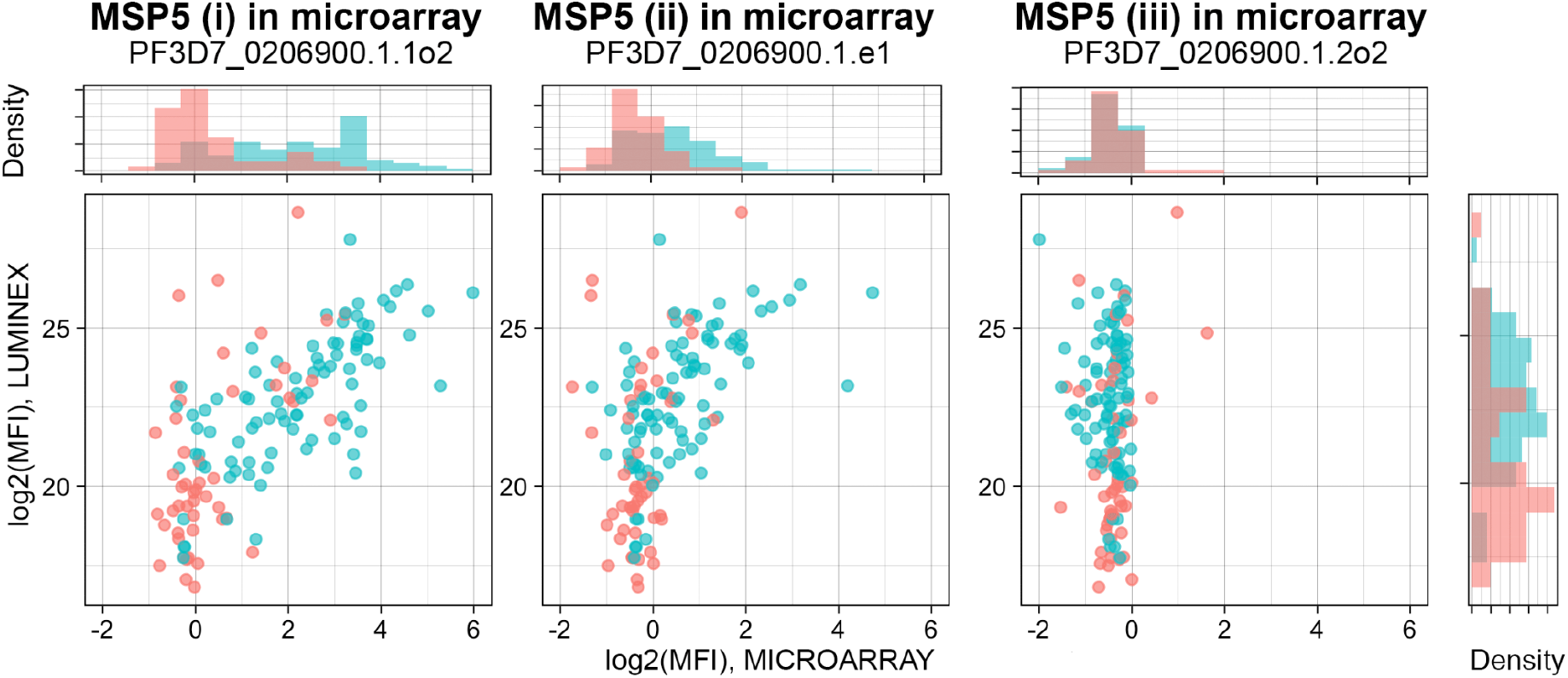
MSP5 Ab levels probed with qSAT (y-axis) against three different constructs of MSP5 probed with a protein array (x-axis). X-axis values are different for each of the three panels (three different protein constructs) but y-axis values do not and produce a single marginal histogram (on the right). Red corresponds to comparator, green to RTS,S vaccinees.

We also see that the marginal plots from Figure 1 do not show a clear bimodality in RTS,S MSP5 Ab levels. This could be due to insufficient sample size, given that we cannot clearly see it in the microarray subset either. On a different note, we also learn that the microarray measurements may not be able to resolve differences at low Ab levels where measurements are packed around zero (background MFI), whereas differences at this low dynamic range can be better resolved with qSAT. The fact that the protein array may poorly resolve low Ab levels and only start measuring at a certain Ab concentrations could also explain why a potential continuous heterogeneity in Ab responses (approximately log-normal in normalized MFI) takes the shape of a bimodal distribution, the first mode being the result of measurement left-censoring at low concentrations, the second mode being the real Ab level order of magnitude peak.

### 4. MSP5 IgGs responses from 813 children from a nested immunological study of a separate phase 2b clinical trial

We measured MSP5 IgG responses from 813 children (age = 12-60 months) vaccinated with either RTS,S/AS02A (n=124) or a comparator vaccine (n= 696) (Alonso et al., 2004).

Again, we estimated the fold increase in the geometric mean of MSP5 Ab MFI levels of RTS,S over comparator vaccinees one month following primary vaccination (2.2 fold-increase, 95% CI [1.8, 2.8], CI obtained with bootstrap N=2000). This estimated fold-increase is lower than those obtained from the protein microarray and qSAT. Disagreement may have several sources since age range, vaccine preparation and MSP5 constructs are different. Furthermore, in Figure 2 log10 MFI measurements are saturated (at about >4.25). This saturation (right censoring) involves underestimating the true expected differences in Ab level means, as a greater number of RTS,S participants with high MSP5 Ab levels had their Ab levels under-measured than comparators. Trajectories in Figure 2 also show that MSP5 Ab that were increased one month after primary vaccination in the RTS,S group later attenuate and are not found 8.5 months after.

**Figure 2.**
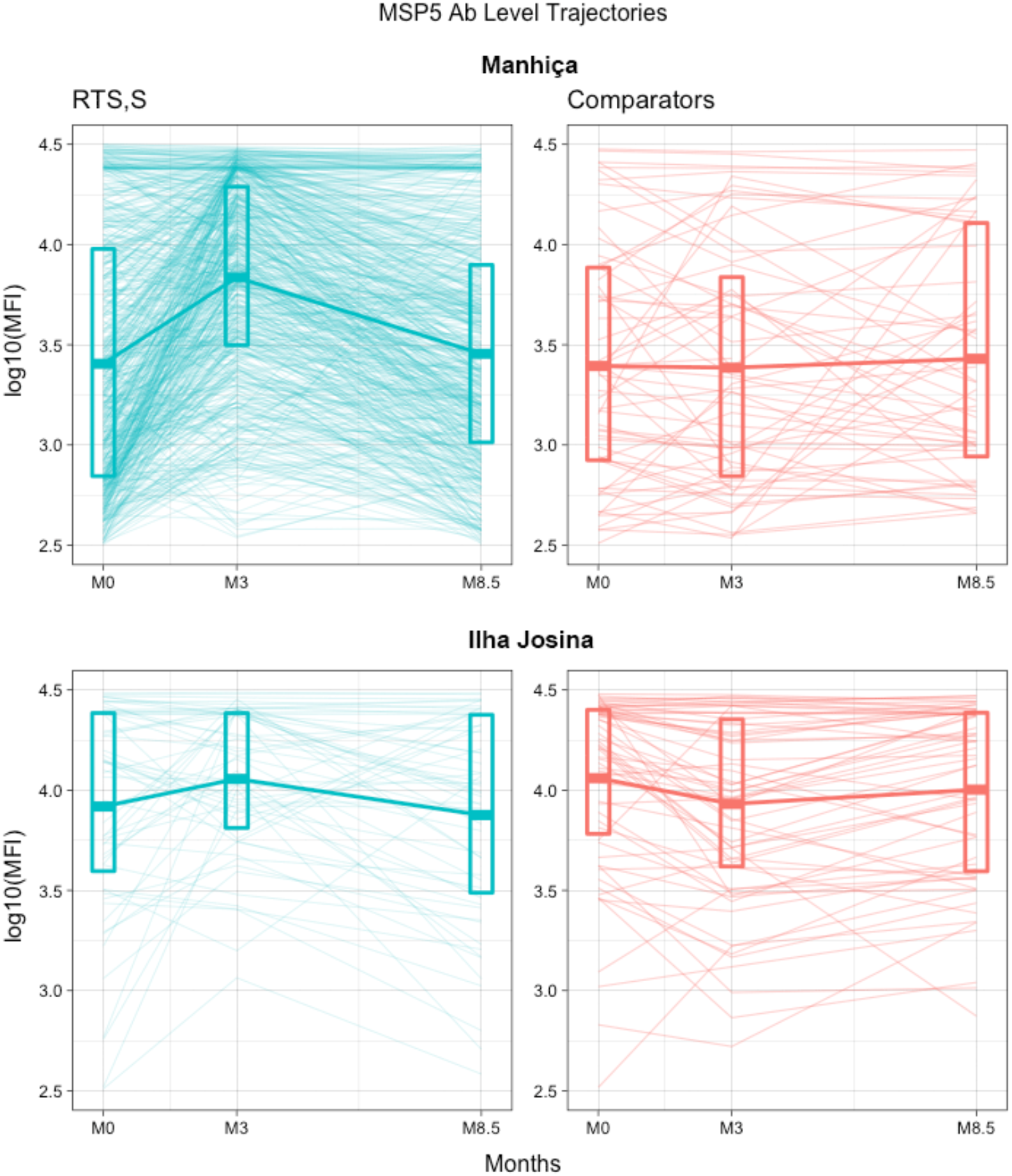
MSP5 Ab level trajectories before primary vaccination (M0), one month after it (M3) and 8.5 months at follow-up (M8.5) for different vaccination groups. *Thin lines on the background correspond to individual trajectories*. Crossbars correspond to group summary statistics per time point with 1^st^, mean and 3^rd^ quartiles represented. TOP: Manhiça cohort representative of low malaria transmission intensity; BOTTOM: Ilha Josina cohort representative of high malaria transmission.

## Supplementary Material 3. Association of vaccine off-target IgG response with NANP repeat and C-terminus CSP IgG concentration and avidity measured by ELISA

To further study the specificity and strength of association between post-vaccination anti-CSP and vaccine off-target IgG levels at M3, antibody (Ab) responses from a subsample of 777 participants (536 RTS,S/AS01E vaccinees) for whom we had ELISA IgG concentration and avidity data against the NANP repeat and C-terminal (C-Term) regions of CSP, were compared to protein array measurements. Details of the ELISA assays and data processing were reported previously (Dobaño et al., 2019).

**Figure 1.**
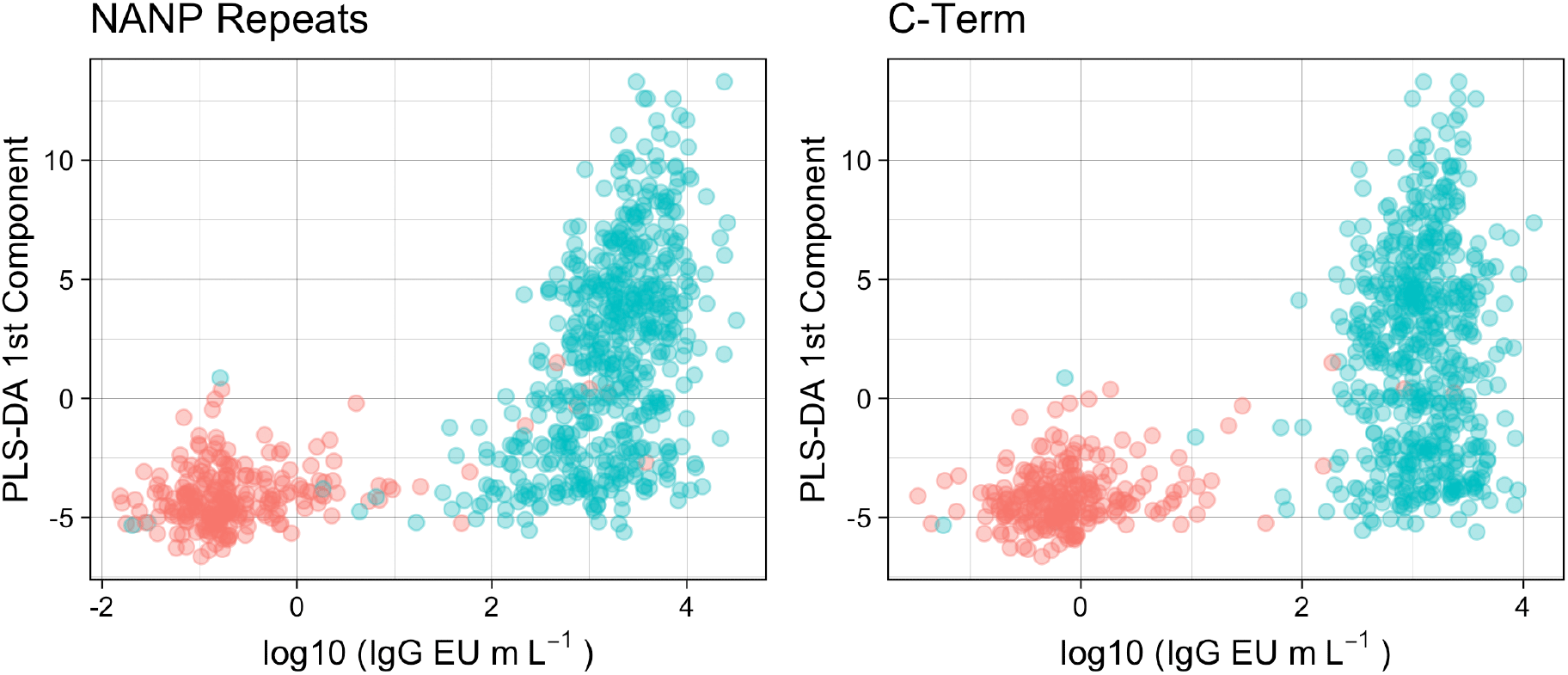
Association of NANP repeat and C-Term CSP IgG concentration with vaccine off-target antibody response. Vaccine off-target antibody response is represented by the 1^st^ PLS-DA component scores (y-axis) of the PLS-DA trained with the log-transformed normalized antibody signals from the protein array other than anti-CSP IgG as predictors and vaccination group as the outcome (see Figure 2 from main manuscript).

***Figure 1*** is a replication of ***Figure 2*** from the main manuscript where CSP Ab levels (x-axis) were now measured by ELISA separately for the NANP repeat and C-Term regions. As noted, high vaccine off-target responses can only occur in RTS,S/AS01E vaccinated individuals (a necessary condition) but within vaccinated individuals a high off-target Ab response is not guaranteed (not a sufficient condition). When focusing on RTS,S/AS01E vaccinated individuals (green dots), a stronger correlation of the off-target Ab response score existed with anti-NANP IgG (ρ=0.48, CI [0.44-0.53], p< 2.2e-16), compared to the one with anti-C-Term IgG (ρ=0.11, CI [0.02, 0.19], p=0.014).

Comparing high against low off-target Ab responders using the dichotomous classification based on the seven most increased off-target Abs, we confirmed an association only with NANP IgG levels in the same direction. RTS,S/AS01E vaccinated individuals classified as high responders had IgG concentrations with a geometric mean 2.19 times higher, 95% CI [1.73, 2.75] (p=1.84e-10), than the low Ab off-target responders. In contrast, no significant difference was found for C-Term IgG levels (fold-difference of 1.31, CI [0.91, 1.32], p=0.84). Fold differences in geometric means were estimated by fitting a multiple regression with log10 IgG as the continuous outcome, the off-target Ab response group as the binary predictor of interest, and adjusting for age group and site.

**Figure 2.**
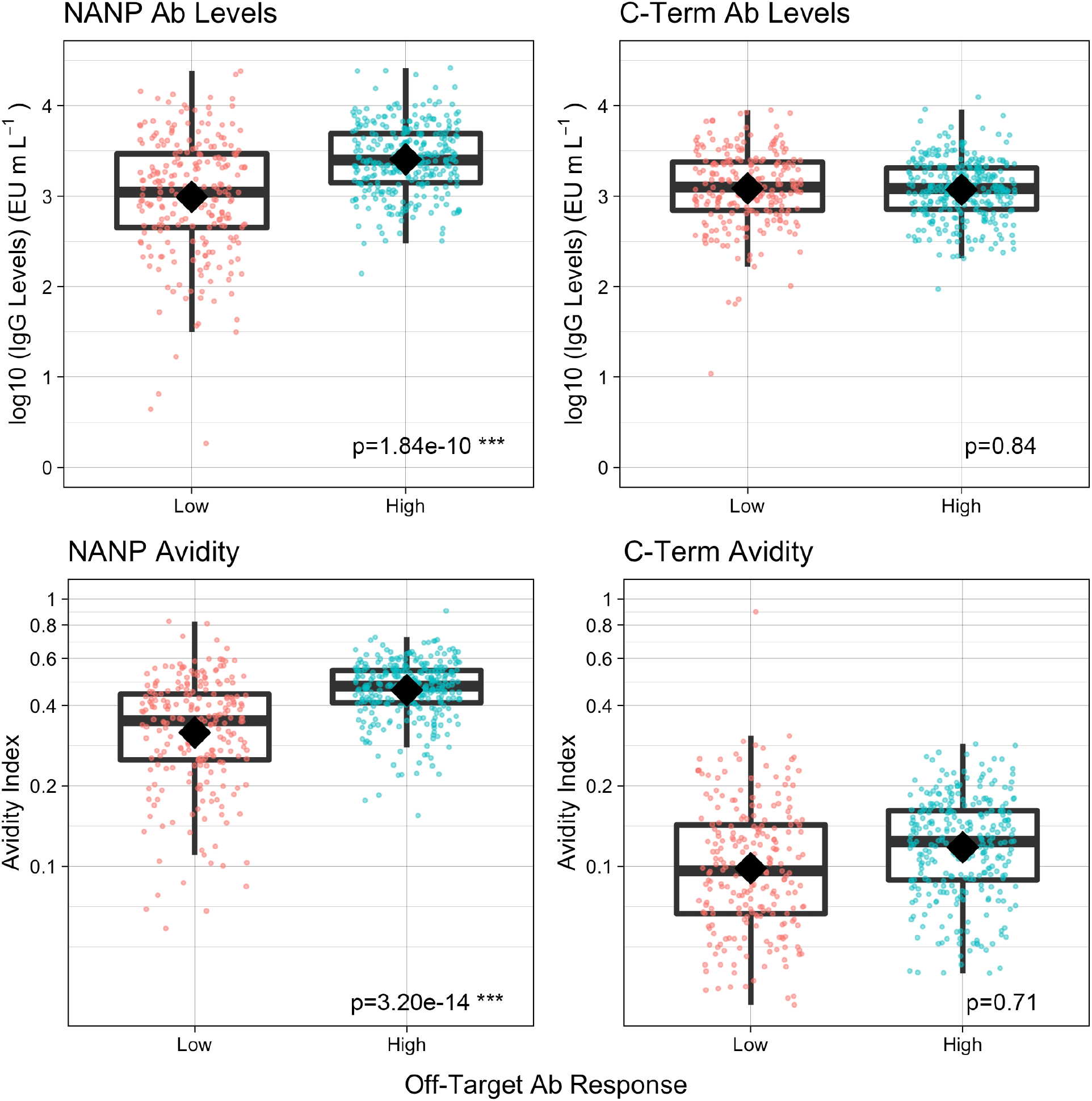
Association of NANP and C-Term CSP Ab levels and avidity with vaccine off-target Ab response group. Boxplots illustrate the medians and the 25^th^ and 75^th^ quartiles, diamonds show the geometric mean, whiskers display the 1.5 interquartile ranges, and coloured dots are measured data. Significance of the geometric mean differences are calculated as t-tests on the regression coefficients in models adjusted by site and age group

Additionally, we conducted the same comparisons using IgG avidity as outcome and also found an association with vaccine off-target Ab response for NANP (a 31% higher geometric mean Avidity Index in high off-target Ab responders, CI [22%, 42%], p=3.20e-14) but not for C-Term (2%, CI [-8%, 13%], p=0.71) (see *Figure 2*). Of note, this significant association of higher post-vaccination NANP IgG avidity in high off-target Ab responders remained when adjusted for NANP IgG levels (28% higher geometric mean Avidity Index, CI [20%, 37%]), considering that Ab avidity correlates with Ab levels (Dobaño et al., 2019).

These results indicate that off-target reactivity is related to the immunogenicity of the immunodominant NANP Abs rather than to that of the subdominant C-Term Abs. Furthermore, if anti-NANP IgGs cross-reacting with other non-vaccine *P. falciparum* antigens in some individuals explain off-target reactivity, then data suggest that these Abs do not bind with less affinity to their cognate NANP target and, therefore, this phenomenon may not be attributed to a potential lack of specificity and/or maturation.

## Supplementary Material 4. Clinical malaria incidence ratios of High over Low off-target antibody responders estimated with and without lost-to-follow-up vaccinees

**Table 1.**
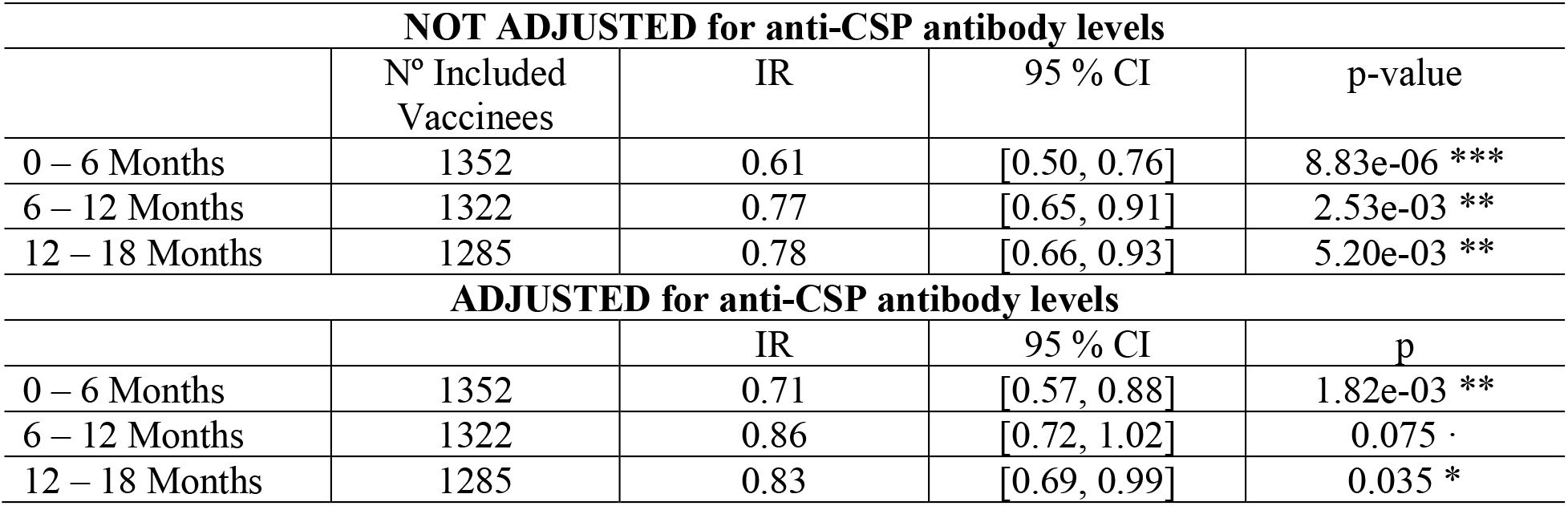
Complete Case Analysis. Individuals who did not complete follow-up (6, 12 or 18 months after M3) were excluded from analyses. These analyses are those reported in the main manuscript

**Table 2.** Incomplete Case Analysis. Individuals who did not complete follow-up (6, 12 or 18 months after M3) were included if followed for longer than 1 month after adjusting for their shorter time at risk in the offset of the negative binomial regression.

| NOT ADJUSTED for anti-CSP antibody levels |  |  |  |  |
| --- | --- | --- | --- | --- |
|  | N° Included Vaccinees | IR | 95 % CI | p-value |
| 0 – 6 Months | 1376 | 0.61 | [0.49, 0.76] | 6.96e-06 *** |
| 6 – 12 Months | 1352 | 0.78 | [0.66, 0.92] | 3.53e-03 ** |
| 12 – 18 Months | 1322 | 0.76 | [0.64, 0.91] | 1.95e-03 ** |
| ADJUSTED for anti-CSP antibody levels |  |  |  |  |
|  |  | IR | 95 % CI | p-value |
| 0 – 6 Months | 1376 | 0.70 | [0.56, 0.87] | 1.21e-03 ** |
| 6 – 12 Months | 1352 | 0.87 | [0.73, 1.03] | 0.10 · |
| 12 – 18 Months | 1322 | 0.80 | [0.67, 0.96] | 0.014 * |

